# Associations of tattooing with health: a population-based cross-sectional study of ∼27,000 US adults

**DOI:** 10.64898/2026.02.23.26346861

**Authors:** Rachel D. McCarty, Britton Trabert, Morgan M. Millar, David Kriebel, Laurie Grieshober, Mollie E. Barnard, Lindsay J. Collin, Jeffrey A. Gilreath, Paul J. Shami, Jennifer A. Doherty

**Author notes:** **Corresponding author:** Rachel D. McCarty, International Agency for Research on Cancer, Rachel D McCarty, 25 avenue Tony Garnier, 69007 Lyon, France.

## Abstract

**Objective:** To characterize associations between tattooing and health status.

**Methods:** We used data from ∼27,000 respondents to the 2020–2022 Utah Behavioral Risk Factor Surveillance System (BRFSS). Multivariable Poisson regression was used to calculate prevalence ratios (PR) and 95% confidence intervals (CI) associating ever receiving a tattoo with physical/mental health status.

**Results:** In this cross-sectional study, ever receiving a tattoo was associated with self-reported “poorer” vs. “excellent” overall health, particularly among women (PR=3.08 [95% CI: 2.26– 4.21]). Tattooing was also associated with obesity (women, PR=1.40 [95% CI: 1.22–1.61]; men, PR=1.21 [95% CI: 1.04–1.40]) and chronic pain (women, PR=1.59 [95% CI: 1.43–1.77]; men, PR=1.55 [95% CI: 1.37–1.76]). Tattooed individuals were more likely to have been diagnosed with a depressive disorder (women, PR=1.64 [95% CI: 1.53–1.75]; men, PR=1.55 [95% CI: 1.39–1.73]) and to have had six or more teeth removed, vs. none (women, PR=2.18 [95% CI: 1.61–2.96]; men, PR=2.88 [95% CI: 2.10–3.95]).

**Conclusions:** Public health entities may consider partnering with tattoo studios and conventions to provide information about nutrition, exercise, dental care, mental health resources, and health screenings.

## Introduction

The United States (US) has one of the highest prevalences of tattooing in the world, with approximately 38% of women and 27% of men having at least one tattoo.^1^ Among adults in Utah, US, tattooed individuals are more likely to be unable to see a doctor due to cost and less likely to receive a flu vaccine than non-tattooed individuals.^2^ Tattooing is also associated with some risky health behaviors, including tobacco smoking^3–5^ and heavy alcohol use.^2,4^ The barriers to healthcare that tattooed individuals may experience, combined with a higher prevalence of risk behaviors suggest that tattooed individuals may experience disproportionate health burdens.

Despite these concerns, tattooed individuals represent a uniquely accessible population for public health interventions. Tattoo studios and conventions serve as community gathering spaces where health promotion campaigns, screenings, and educational resources can effectively reach this population. Understanding the health profile of tattooed individuals is therefore critical for developing targeted interventions in these settings. In this study, we aimed to quantify associations between tattooing and health status, including self-reported physical and mental health outcomes in a population-representative sample of ∼27,000 Utah adults.

## Methods

We used a cross-sectional study design including respondents to the 2020, 2021, and 2022 Utah Behavioral Risk Factor Surveillance System (BRFSS) surveys. The BRFSS is a population-based telephone health survey administered throughout the US. The BRFSS uses a disproportionate stratified sampling design which stratifies by phone type and region.^7^ Design weights and iterative proportional fitting (raking) ensure the sample is reflective of the underlying population. The methodology used in this study has been described previously.^2^ Participants to the 2020, 2021, and 2022 Utah BRFSS surveys were asked about any tattoos they had received with a tattoo machine.

The health status variables included in our analyses were obtained from the same BRFSS surveys and include: overall health (excellent, very good, good, fair, poor); number of days during the past 30 days when physical health was not good; average hours of sleep per night (<7, 7–9, >9); body mass index (BMI); physical activity in the past 30 days (yes/no); chronic pain (yes/no), prescription opioid use for chronic pain (yes/no), visited a dentist in the past year (yes/no); number of permanent teeth removed due to tooth decay or gum disease; ever had a depressive disorder (yes/no); number of days during the past 30 days when mental health was not good; and stress during the past 30 days (never, rarely, sometimes, usually, always).

The chronic health condition variables included in supplemental analyses were ever diagnosis of the following conditions: asthma (current and ever); chronic obstructive pulmonary disease (COPD)/emphysema/chronic bronchitis; stroke; heart attack; angina or coronary heart disease; high blood pressure; high cholesterol; kidney disease; diabetes; arthritis/rheumatoid arthritis/gout/lupus/fibromyalgia; and any cancer.

We calculated prevalence ratios (PRs) and 95% confidence intervals (CIs) associating ever receiving a tattoo (yes/no) with each health status variable, stratified by sex. We fit modified Poisson regression models for binary health status variables and multinomial models for categorical health status variables with more than two categories. The use of robust (sandwich) variance estimators provides valid standard errors regardless of the underlying variance structure, addressing concerns about overdispersion that can arise with Poisson models. Models were adjusted for age (continuous) and education level (less than high school, high school diploma, some college, college graduate or more). We assessed multicollinearity between age and education through model comparisons and cross-tabulation analyses, confirming both variables were necessary to avoid confounding bias (Supplementary Table 1 for model comparisons). We then fit models further adjusted for ever tobacco smoking (yes/no), heavy drinking (yes/no), BMI (<18.5 kg/m^2^, 18.5–24.9, 25.0–29.9, 30+), and physical activity in the past 30 days (yes/no) to examine the impact of these variables on the observed associations. The number of individuals reporting specific cancer types was small, so we further fit a multinomial model associating ever receiving a tattoo with cancer type among all individuals, controlling for sex. In supplemental analyses to assess potential residual confounding by tobacco smoking status, we fit comparable models restricted to individuals who reported never smoking.

All models incorporated the BRFSS design weights to account for the complex survey design; while the reported sample sizes are unweighted, the percentages are weighted and therefore more accurately reflect the underlying Utah population. We assessed model diagnostics including model convergence, sample sizes across exposure-outcome categories (minimum cell size = 5), and variance inflation factors (VIFs). Analyses were conducted with R Statistical Software (v4.3.1; R core team 2023; Vienna, Austria).

## Results

The majority of participants reported non-Hispanic White race (78% of women and 77% of men), and 13% reported Hispanic ethnicity (Table 1). The age distribution was similar for women and men. The largest group was those aged 60 or older (26% of women, 24% of men), followed by those aged 30–39 (19% for both women and men). Most participants were married (∼60%) and had attained more than a high school education (70% of women, 67% of men). The prevalence of tattooing was 24% overall, with 26% of women and 22% of men reporting having at least one tattoo.

**Table 1.** Demographics of BRFSS respondents and corresponding tattoo prevalences in women and men.

|  | Women<br>(n=13,445) |  |  | Men<br>(n=13,178) |  |  |
| --- | --- | --- | --- | --- | --- | --- |
|  | n | (%) | Tattoo<br>prevalence | n | (%) | Tattoo<br>prevalence |
| **Overall** |  |  | 26% |  |  | 22% |
| **Survey years** |  |  |  |  |  |  |
| 2020 | 4,804 | (33%) | 26% | 4,569 | (33%) | 21% |
| 2021 | 4,665 | (35%) | 26% | 4,669 | (35%) | 22% |
| 2022 | 3,976 | (32%) | 26% | 3,960 | (33%) | 21% |
| **Race and ethnicity** |  |  |  |  |  |  |
| Hispanic | 1,241 | (13%) | 30% | 1,146 | (13%) | 31% |
| Non-Hispanic American Indian/Alaskan |  |  |  |  |  |  |
| Native | 136 | (1.3%) | 47% | 99 | (0.9%) | 40% |
| Non-Hispanic Asian | 141 | (2.3%) | 16% | 161 | (2.7%) | 16% |
| Non-Hispanic Black | 68 | (0.8%) | 24% | 97 | (1.2%) | 18% |
| Non-Hispanic Multiracial | 143 | (1.6%) | 37% | 147 | (1.7%) | 40% |
| Non-Hispanic Pacific Islander | 53 | (0.8%) | 41% | 57 | (0.9%) | 34% |
| Non-Hispanic White | 11,460 | (78%) | 25% | 11,089 | (77%) | 20% |
| Non-Hispanic Other | 17 | (<0.1%) | 1% | 42 | (0.4%) | 19% |
| Unknown/missing | 186 | (1.3%) | 26% | 340 | (2.4%) | 20% |
| **Age** |  |  |  |  |  |  |
| 18-24 | 928 | (14%) | 34% | 1,264 | (16%) | 22% |
| 25-29 | 840 | (9.3%) | 43% | 885 | (9.3%) | 33% |
| 30-39 | 1,933 | (19%) | 39% | 2,011 | (19%) | 31% |
| 40-49 | 2,286 | (17%) | 27% | 2,150 | (17%) | 27% |
| 50-59 | 1,883 | (13%) | 20% | 1,888 | (13%) | 17% |
| 60 or older | 5,408 | (26%) | 9% | 4,807 | (24%) | 9% |
| Unknown/missing | 167 | (1.1%) | 10% | 173 | (1.2%) | 7% |
| **Marital status** |  |  |  |  |  |  |
| Married | 8,356 | (59%) | 21% | 8,881 | (60%) | 18% |
| Not married | 5,033 | (40%) | 33% | 4,205 | (39%) | 28% |
| Unknown/missing | 56 | (0.4%) | 16% | 92 | (0.7%) | 24% |
| **Education** |  |  |  |  |  |  |
| <High school diploma | 565 | (7.6%) | 29% | 567 | (7.8%) | 33% |
| High school diploma or GED | 3,198 | (23%) | 34% | 3,066 | (25%) | 30% |
| Some college | 4,583 | (41%) | 27% | 3,737 | (35%) | 23% |
| College graduate or more | 5,073 | (29%) | 18% | 5,771 | (32%) | 11% |
| Unknown/missing | 26 | (0.2%) | 4% | 37 | (0.3%) | 30% |
| **Ever smoked tobacco** |  |  |  |  |  |  |
| Yes | 2,666 | (19%) | 54% | 3,748 | (28%) | 43% |
| No | 10,737 | (81%) | 20% | 9,368 | (71%) | 13% |
| Unknown/missing | 42 | (0.3%) | 38% | 62 | (0.4%) | 23% |
| **Heavy drinking in past month** |  |  |  |  |  |  |
| Yes | 461 | (3.9%) | 62% | 556 | (4.8%) | 49% |
| No | 12,852 | (95%) | 24% | 12,448 | (94%) | 20% |
| Unknown/missing | 132 | (1.1%) | 38% | 174 | (1.5%) | 33% |
| **Latter-day Saints religion** |  |  |  |  |  |  |
| Yes | 7,745 | (52%) | 10% | 7,223 | (48%) | 9% |
| No | 5,510 | (47%) | 44% | 5,749 | (51%) | 34% |
| Unknown/missing | 190 | (1.3%) | 20% | 206 | (1.5%) | 18% |

Note: n's are unweighted. Percentages are weighted to account for the complex survey design of the BRFSS. Incorporating these survey weights allows the estimates to more accurately reflect the underlying Utah population.
The prevalence of tattooing was 24% overall
Abbreviation: General Education Development (GED)

For the results below, we confirmed that: (1) all models converged successfully, (2) sample sizes were adequate across all exposure-outcome categories (minimum cell size = 5), and (3) variance inflation factors (VIFs < 2.5) indicated no problematic multicollinearity among covariates.

### Health status: physical health

Among women, those with tattoos were more likely to report poorer overall health status compared with those without tattoos, after adjustment for age and education level. Specifically, the prevalence of reporting “fair” versus “excellent” health was over 1.6 times higher (PR=1.66 [95% CI: 1.34–2.06]) and the prevalence of reporting “poor” versus “excellent” health was 3 times higher (PR=3.08 [95% CI: 2.26–4.21]) compared to women without tattoos (Table 2). Among men, patterns were weaker but with similar trends. Women with tattoos were about 1.3 times more likely (PR=1.33 [95% CI: 1.03–1.72]) and men with tattoos were 1.6 times more likely (PR=1.60 [95% CI: 1.18–2.16]) to report 8–14 days (versus none) when their physical health was not good during the past 30 days compared with those without tattoos. Women with tattoos were about 1.7 times more likely (PR=1.69 [95% CI: 1.42–2.02]) and men with tattoos were about 1.5 times more likely (PR=1.45 [95% CI: 1.17–1.80]) to report 15 or more days with poor physical health than those without tattoos. Associations with overall health were still two times higher for tattooed women after additionally controlling for having ever smoked tobacco, heavy drinking, BMI, and physical activity, but associations were attenuated for men.

**Table 2.** Associations between ever receiving a tattoo and physical, oral, and mental health status among women and men.

|  | Women |  |  |  |  |  | Men |  |  |  |  |  |
| --- | --- | --- | --- | --- | --- | --- | --- | --- | --- | --- | --- | --- |
|  | Never tattooed<br>(n=10,599) |  | Ever tattooed<br>(n=2,846) |  | Model 1 | Model 2 | Never tattooed<br>(n=10,810) |  | Ever tattooed<br>(n=2,368) |  | Model 1 | Model 2 |
|  | n | (%) | n | (%) | PR (95% CI) | PR (95% CI) | n | (%) | n | (%) | PR (95% CI) | PR (95% CI) |
| **Physical health** |  |  |  |  |  |  |  |  |  |  |  |  |
| **Overall health** |  |  |  |  |  |  |  |  |  |  |  |  |
| Excellent | 2,146 | (22%) | 521 | (18%) | Ref | Ref | 2,392 | (24%) | 483 | (22%) | Ref | Ref |
| Very good | 3,983 | (38%) | 931 | (32%) | 1.12 (0.96, 1.32) | 1.00 (0.84, 1.19) | 3,885 | (36%) | 744 | (32%) | 1.03 (0.88, 1.21) | 0.96 (0.81, 1.14) |
| Good | 2,992 | (27%) | 899 | (33%) | 1.61 (1.36, 1.89) | 1.28 (1.06, 1.55) | 3,184 | (29%) | 762 | (32%) | 1.25 (1.06, 1.46) | 1.03 (0.86, 1.23) |
| Fair | 1,102 | (10%) | 358 | (12%) | 1.66 (1.34, 2.06) | 1.31 (1.01, 1.70) | 986 | (8.6%) | 288 | (12%) | 1.56 (1.24, 1.96) | 1.10 (0.85, 1.42) |
| Poor | 349 | (2.5%) | 130 | (4.2%) | 3.08 (2.26, 4.21) | 2.20 (1.55, 3.12) | 341 | (2.6%) | 85 | (2.7%) | 1.41 (0.98, 2.03) | 0.99 (0.67, 1.45) |
| Unknown/missing | 27 | (0.3%) | * | (0.2%) |  |  | 22 | (0.2%) | * | (0.2%) |  |  |
| **Number of days during the past 30 days when physical health was not good** |  |  |  |  |  |  |  |  |  |  |  |  |
| None | 6,286 | (58%) | 1,569 | (54%) | Ref | Ref | 7,259 | (67%) | 1,535 | (65%) | Ref | Ref |
| 1-7 | 2,451 | (25%) | 661 | (24%) | 0.89 (0.78, 1.02) | 0.83 (0.71, 0.96) | 2,180 | (22%) | 463 | (20%) | 0.92 (0.80, 1.07) | 0.91 (0.77, 1.07) |
| 8-14 | 427 | (4.4%) | 161 | (6.0%) | 1.33 (1.03, 1.72) | 1.31 (0.99, 1.73) | 324 | (2.9%) | 94 | (4.2%) | 1.60 (1.18, 2.16) | 1.46 (1.06, 2.02) |
| 15+ | 1,167 | (9.8%) | 408 | (14%) | 1.69 (1.42, 2.02) | 1.38 (1.13, 1.70) | 869 | (6.9%) | 233 | (8.5%) | 1.45 (1.17, 1.80) | 1.25 (0.99, 1.58) |
| Unknown/missing | 268 | (2.4%) | 47 | (1.5%) |  |  | 178 | (1.4%) | 43 | (1.8%) |  |  |
| **Average hours of sleep per night (2020 and 2022)<sup>a</sup>** |  |  |  |  |  |  |  |  |  |  |  |  |
| <7 | 1,827 | (28%) | 711 | (39%) | 1.58 (1.36, 1.83) | 1.44 (1.22, 1.70) | 2,049 | (31%) | 633 | (44%) | 1.64 (1.41, 1.90) | 1.49 (1.26, 1.76) |
| 7-9 | 4,747 | (68%) | 1,057 | (55%) | Ref | Ref | 4,751 | (66%) | 799 | (53%) | Ref | Ref |
| >9 | 251 | (3.1%) | 79 | (4.1%) | 1.83 (1.26, 2.65) | 1.79 (1.18, 2.71) | 163 | (2.0%) | 41 | (2.4%) | 1.44 (0.91, 2.26) | 1.11 (0.67, 1.83) |
| Unknown/missing | 86 | (1.3%) | 22 | (1.3%) |  |  | 53 | (0.7%) | 20 | (0.7%) |  |  |
| **Body mass index (kg/m<sup>2</sup>)** |  |  |  |  |  |  |  |  |  |  |  |  |
| Underweight (<18.5) | 195 | (2.0%) | 52 | (2.3%) | 1.08 (0.70, 1.66) | 1.02 (0.65, 1.60) | 134 | (1.6%) | 33 | (1.6%) | 0.71 (0.44, 1.14) | 0.74 (0.45, 1.20) |
| Normal weight (18.5-24.9) | 3,487 | (34%) | 920 | (32%) | Ref | Ref<sup>b</sup> | 2,736 | (28%) | 638 | (29%) | Ref | Ref<sup>b</sup> |
| Overweight (25.0-29.9) | 2,881 | (25%) | 719 | (25%) | 1.17 (1.01, 1.36) | 1.14 (0.97, 1.33) | 4,310 | (37%) | 877 | (36%) | 1.12 (0.97, 1.30) | 1.10 (0.95, 1.29) |
| Obese (30+) | 2,772 | (26%) | 903 | (32%) | 1.40 (1.22, 1.61) | 1.36 (1.16, 1.58) | 3,271 | (29%) | 777 | (32%) | 1.21 (1.04, 1.40) | 1.23 (1.05, 1.44) |
| Unknown/missing | 1,264 | (12%) | 252 | (9.0%) |  |  | 359 | (3.9%) | 43 | (1.9%) |  |  |
| **Physical activity in past 30 days** |  |  |  |  |  |  |  |  |  |  |  |  |
| Yes | 8,504 | (82%) | 2,236 | (80%) | Ref | Ref<sup>c</sup> | 9,197 | (86%) | 1,946 | (83%) | Ref | Ref<sup>c</sup> |
| No | 2,080 | (18%) | 608 | (20%) | 1.21 (1.09, 1.36) | 1.10 (0.97, 1.25) | 1,590 | (14%) | 417 | (16%) | 1.09 (0.95, 1.24) | 1.01 (0.88, 1.17) |
| Unknown/missing | 15 | (0.1%) | * | (<0.1%) |  |  | 23 | (0.2%) | * | (0.2%) |  |  |
| **Chronic pain (2020 and 2022)<sup>d</sup>** |  |  |  |  |  |  |  |  |  |  |  |  |
| No | 3,538 | (69%) | 833 | (59%) | Ref | Ref | 3,825 | (74%) | 696 | (64%) | Ref | Ref |
| Yes | 1,668 | (28%) | 570 | (37%) | 1.59 (1.43, 1.77) | 1.39 (1.23, 1.56) | 1,347 | (23%) | 436 | (33%) | 1.55 (1.37, 1.76) | 1.27 (1.11, 1.46) |
| Unknown/missing | 151 | (3.2%) | 49 | (3.9%) |  |  | 157 | (3.0%) | 32 | (3.0%) |  |  |
| **Prescription opioid use for chronic pain (2020 and 2022)<sup>d</sup>** |  |  |  |  |  |  |  |  |  |  |  |  |
| No | 1,350 | (83%) | 422 | (78%) | Ref | Ref | 1,089 | (83%) | 371 | (88%) | Ref | Ref |
| Yes | 305 | (16%) | 141 | (20%) | 1.62 (1.23, 2.12) | 1.53 (1.14, 2.04) | 244 | (16%) | 63 | (12%) | 0.91 (0.65, 1.27) | 0.82 (0.58, 1.16) |
| Unknown/missing | 13 | (0.6%) | * | (1.7%) |  |  | 14 | (0.7%) | * | (0.3%) |  |  |
| **Oral health** |  |  |  |  |  |  |  |  |  |  |  |  |
| **Visited a dentist in the past year (2020 and 2022)** |  |  |  |  |  |  |  |  |  |  |  |  |
| Yes | 5,369 | (76%) | 1,266 | (69%) | Ref | Ref | 5,109 | (72%) | 905 | (61%) | Ref | Ref |
| No | 1,480 | (23%) | 589 | (31%) | 1.21 (1.09, 1.35) | 1.04 (0.92, 1.18) | 1,850 | (28%) | 576 | (38%) | 1.20 (1.09, 1.33) | 1.06 (0.95, 1.18) |
| Unknown/missing | 62 | (0.9%) | 14 | (0.7%) |  |  | 57 | (0.7%) | 12 | (0.6%) |  |  |
| **Number of permanent teeth removed due to tooth decay or gum disease (2020 and 2022)** |  |  |  |  |  |  |  |  |  |  |  |  |
| None | 4,099 | (66%) | 1,117 | (64%) | Ref | Ref | 4,203 | (67%) | 828 | (60%) | Ref | Ref |
| 1-5 | 1,902 | (24%) | 506 | (26%) | 1.52 (1.29, 1.80) | 1.33 (1.10, 1.61) | 1,967 | (24%) | 448 | (28%) | 1.66 (1.40, 1.98) | 1.35 (1.11, 1.65) |
| 6 or more but not all | 482 | (5.5%) | 128 | (5.3%) | 2.18 (1.61, 2.96) | 1.48 (1.03, 2.12) | 450 | (4.9%) | 133 | (7.8%) | 2.88 (2.10, 3.95) | 1.80 (1.27, 2.56) |
| All | 280 | (3.1%) | 97 | (3.3%) | 2.99 (1.93, 4.61) | 1.97 (1.17, 3.31) | 276 | (3.0%) | 70 | (3.0%) | 2.16 (1.43, 3.26) | 1.11 (0.72, 1.71) |
| Unknown/missing | 148 | (1.7%) | 21 | (0.9%) |  |  | 120 | (1.3%) | 14 | (0.7%) |  |  |
| **Mental health** |  |  |  |  |  |  |  |  |  |  |  |  |
| **Ever had a depressive disorder** |  |  |  |  |  |  |  |  |  |  |  |  |
| No | 7,858 | (72%) | 1,476 | (50%) | Ref | Ref | 9,202 | (84%) | 1,740 | (74%) | Ref | Ref |
| Yes | 2,677 | (27%) | 1,352 | (49%) | 1.64 (1.53, 1.75) | 1.41 (1.30, 1.52) | 1,538 | (15%) | 612 | (26%) | 1.55 (1.39, 1.73) | 1.34 (1.18, 1.50) |
| Unknown/missing | 64 | (0.7%) | 18 | (0.8%) |  |  | 70 | (0.8%) | 16 | (0.6%) |  |  |
| **Number of days during the past 30 days when mental health was not good** |  |  |  |  |  |  |  |  |  |  |  |  |
| None | 5,472 | (45%) | 943 | (30%) | Ref | Ref | 7,041 | (60%) | 1,259 | (49%) | Ref | Ref |
| 1-7 | 2,978 | (31%) | 787 | (28%) | 1.08 (0.94, 1.25) | 0.99 (0.85, 1.16) | 2,279 | (24%) | 504 | (24%) | 1.09 (0.94, 1.27) | 1.12 (0.96, 1.32) |
| 8-14 | 632 | (7.3%) | 279 | (10%) | 1.51 (1.22, 1.86) | 1.35 (1.07, 1.70) | 420 | (4.7%) | 148 | (5.9%) | 1.23 (0.95, 1.59) | 1.11 (0.82, 1.50) |
| 15+ | 1,319 | (15%) | 798 | (31%) | 2.21 (1.91, 2.57) | 1.71 (1.44, 2.03) | 920 | (9.8%) | 421 | (19%) | 1.88 (1.59, 2.23) | 1.53 (1.27, 1.84) |
| Unknown/missing | 198 | (2.1%) | 39 | (1.5%) |  |  | 150 | (1.6%) | 36 | (1.1%) |  |  |
| **Within the last 30 days, how often have you felt stress?<sup>e</sup> (2022)** |  |  |  |  |  |  |  |  |  |  |  |  |
| Never | 926 | (26%) | 166 | (19%) | Ref | Ref | 1,319 | (37%) | 212 | (27%) | Ref | Ref |
| Rarely | 1,056 | (32%) | 223 | (25%) | 0.91 (0.67, 1.22) | 0.85 (0.61, 1.18) | 1,017 | (32%) | 180 | (26%) | 1.06 (0.80, 1.40) | 1.06 (0.79, 1.43) |
| Sometimes | 770 | (29%) | 271 | (30%) | 1.03 (0.77, 1.38) | 0.97 (0.69, 1.35) | 611 | (20%) | 164 | (27%) | 1.72 (1.28, 2.30) | 1.52 (1.09, 2.13) |
| Usually | 247 | (9.4%) | 117 | (14%) | 1.29 (0.88, 1.87) | 1.05 (0.70, 1.58) | 216 | (8.2%) | 84 | (12%) | 1.67 (1.13, 2.47) | 1.18 (0.76, 1.84) |
| Always | 102 | (4.0%) | 87 | (11%) | 2.42 (1.54, 3.79) | 1.81 (1.05, 3.12) | 83 | (3.0%) | 62 | (7.9%) | 2.97 (1.87, 4.72) | 2.20 (1.31, 3.71) |
| Unknown/missing | * | (0.2%) | * | (0.3%) |  |  | 11 | (0.3%) | * | (0.1%) |  |  |

Note: n's are unweighted. Percentages are weighted and models incorporate the BRFSS weights to account for the complex survey design. Model 1 is adjusted for age and education level. Model 2 is adjusted for age, education level, ever tobacco smoking, heavy drinking, BMI, and physical activity in the past 30 days.
\*Censored due to cell sizes <11
<sup>a</sup>Year indicators in parentheses denote the survey years in which that variable was assessed. Variables without year indicators were included in all three survey years.
<sup>b</sup>BMI not included as adjustment variables in model 2
<sup>c</sup>Physical activity not included as an adjustment variable in model 2
<sup>d</sup>2020: All participants (both survey legs); 2022: 50% of participants (one survey leg)
<sup>e</sup>Stress means a situation in which a person feels tense, restless, nervous or anxious or is unable to sleep at night because their mind is troubled all the time. Within the last 30 days how often have you felt this kind of stress?

Tattooing was also associated with both reporting less than 7 hours and reporting more than 9 hours of sleep per night. Women and men with tattoos were about 1.6 times more likely to sleep less than 7 hours per night (women: PR=1.58 [95% CI: 1.36–1.83]; men: PR=1.64 [95% CI: 1.41–1.90]) compared with those without tattoos. Tattooed women were 1.8 times more likely (PR=1.83 [95% CI: 1.26–2.65]) and tattooed men were 1.4 times more likely (PR=1.44 [95% CI 0.91–2.26]) to sleep 9 or more hours per night compared with non-tattooed individuals. Tattooed women were 1.4 times (PR=1.40 [95% CI: 1.22–1.61]) and tattooed men were about 1.2 times more likely (PR=1.21 [95% CI: 1.04–1.40]) to be obese (BMI 30+) compared with non-tattooed individuals. Tattooed women (but not men) were about 1.2 times more likely to report no physical activity in the past 30 days (PR=1.21 [95% CI 1.09–1.36]). Tattooed individuals were roughly 1.6 times more likely to report chronic pain than non-tattooed (women: PR=1.59 [95% CI 1.43–1.77]; men: PR=1.55 [95% CI 1.37–1.76]). Tattooed women (but not men) were about 1.6 times more likely to use a prescribed opioid for chronic pain than non-tattooed individuals (PR=1.62 [95% CI 1.23–2.12]). Tattooed individuals were about 1.2 times less likely to have seen a dentist in the past year than non-tattooed individuals (women: PR=1.21 [95% CI: 1.09– 1.35; men: PR=1.20 [95% CI: 1.09–1.33]). Tattooed women and men were about 2–3 times more likely to have had six or more permanent teeth removed due to tooth decay or gum disease than non-tattooed individuals (women: PR=2.18 [95% CI: 1.61–2.96]; men: PR=2.88 [95% CI: 2.10–3.95]). In general, associations were attenuated after further consideration of having ever smoked tobacco, heavy drinking, BMI, and physical activity.

### Health status: mental health

Being tattooed was associated with depressive disorders, poor mental health, and stress. Tattooed individuals were about 1.6 times more likely to have ever been diagnosed with a depressive disorder compared with non-tattooed individuals (women: PR=1.64 [95% CI: 1.53– 1.75; men: PR=1.55 [95% CI: 1.39–1.73]). Individuals with tattoos were also more likely to report more days with poor mental health during the past 30 days compared with those without tattoos. Tattooed women were over 2 times more likely (PR=2.21 [95% CI: 1.91–2.57]) and tattooed men were about 1.9 times more likely (PR=1.88 [95% CI: 1.59–2.23]) to report 15 or more poor mental health days compared with those without tattoos. Tattooed women were over two-times more likely and tattooed men were nearly three-times more likely to report always experiencing stress in the last 30 days, defined as feeling tense, restless, nervous, or anxious, or unable to sleep at night compared with non-tattooed individuals (women: PR=2.42 [95% CI: 1.54–3.79]; men: PR=2.97 [95% CI: 1.87–4.72]).

### Chronic health conditions

Ever receiving a tattoo was associated with an increased prevalence of several chronic health conditions compared with people who were never tattooed (Supplementary Table 2). Among both women and men, tattooing was associated with asthma (women: PR=1.37 [95% CI 1.21–1.55]; men: PR=1.26 [95% CI 1.10–1.45]) and arthritis, rheumatoid arthritis, gout, lupus, or fibromyalgia (women: PR=1.20 [95% CI 1.09–1.33]; men: PR=1.20 [95% CI 1.06–1.35]). Among men, tattooing was associated with stroke (PR=1.77 [95% CI 1.26–2.50]). Among women, tattooing was associated with kidney disease (PR=1.44 [95% CI 1.03–2.02]), breast cancer (PR=1.87 [95% CI 1.25–2.80]), and cervical cancer (PR=2.44 [95% CI 1.32–4.49]). Tattooed women and men combined were also more likely to have had colorectal cancer (PR=1.42 [95% CI 0.78–2.61]) and bladder cancer (PR=2.00 [95% CI 0.85–4.68]) (Supplementary Table 3), though these estimates were imprecise as they were based on only 20 tattooed individuals with colorectal cancer and 11 individuals with bladder cancer.

Models restricted to never smokers produced similar effect estimates to overall findings (Supplementary Tables 4-6). Among women and men, the overall associations and the associations among never smokers were quite similar (Table 2 and Supplemental Table 4). For some variables such as number of permanent teeth removed due to tooth decay or gum disease, associations were attenuated among never smokers, however for other variables such as how often they felt stress, the associations were stronger when limited to never smokers.

For chronic conditions, restricting to never smokers generally produced similar or stronger effect estimates rather than attenuating them, suggesting the tattoo-health associations are not primarily explained by confounding from smoking (Supp Table 2 and Supp Table 5).

The cancer associations that were most robust in the overall population (breast and gynecologic in women) persist or strengthen when restricted to never smokers (Supp Table 3 and Supp Table 6).

## Discussion

In this contemporary, cross-sectional study of tattooing and health status in ∼27,000 individuals, we observed that tattooing was associated with several physical and mental health conditions. These include poorer overall physical health, ever diagnosis of a depressive disorder, more days during the past 30 with poor mental health, inadequate or excessive sleep, stress, permanent tooth loss, chronic pain and asthma. Since tattooing and chronic conditions were assessed simultaneously, we cannot establish whether tattoos preceded or followed the health events. In some cases, the tattoo was likely obtained after the health event. For example, among women with breast cancer, some of the tattooed individuals may have received their tattoos following cancer treatment if they received reconstructive surgery with nipple tattooing. While biological mechanisms linking tattoo ink components to health outcomes cannot be excluded, tattooing is also strongly associated with smoking and other risk-taking behaviors that themselves predict poor health outcomes, making it difficult to disentangle these relationships without temporal data. Therefore, these associations should be interpreted as descriptive rather than causal.

Our findings are consistent with a prior online survey conducted in 2016 of ∼2,000 US adults using Amazon’s Mechanical Turk platform, which observed a strong relationship between tattooing and ever receiving a mental health condition diagnosis, as well as difficulty sleeping.^3^ While that study did not observe associations between tattooing and overall health status,^3^ another online survey of ∼450 US adults conducted in 2008 found that participants with a tattoo were more likely to have used a sick day from work in the past 30 days than individuals without a tattoo.^6^ Our study is the first to report on permanent tooth loss, chronic pain, asthma, breast cancer, and gynecologic cancers.

Strengths of this study include the large population-based design, and the ability to examine associations between tattooing and a wide variety of health outcomes. Additionally, missing data were minimal for most variables, and the proportion of missing data was comparable between ever and never tattooed individuals. However, the cross-sectional design prohibits the ability to examine temporality between tattoo exposure and development of health outcomes, as the BRFSS does not collect data on the timing of medical diagnoses. Further because exposure and outcome were measured simultaneously, both are subject to potential misclassification. Our reliance on self-reported medical diagnoses is an important limitation which could lead to misclassification. If tattooed individuals face greater barriers to healthcare access (as prior literature suggests), they may be less likely to receive formal diagnoses, potentially biasing our estimates toward the null and underestimating true associations. Conversely, differential reporting accuracy could bias estimates in either direction. This study may also be affected by selection bias as non-respondents may have been less healthy than respondents. Despite these limitations, to our knowledge, this is the largest population-based study to date which describes associations between health status and tattooing prevalence, providing justification for future studies to evaluate potential causal relationships.

## Conclusions

These findings suggest that tattooing is correlated with several poor physical and mental health outcomes. Given that individuals with tattoos face documented barriers to healthcare access, tattoo conventions and studios present opportunities for partnerships to reach individuals with health promotion activities, disease screening programs, and linkage to care services. Such venues may be particularly well-suited for interventions targeting modifiable risk factors including tobacco use, alcohol consumption, and physical inactivity.

## Data Availability

The 2020 and 2021 BRFSS data used in this study are available from the Utah Department of Health and Human Services. Restrictions apply to the availability of these data.

## Declarations

### Ethics approval and consent to participant

No approval was needed for this study. All analyses utilized secondary de-identified data from the BRFSS survey.

### Consent for publication

Not applicable

### Availability of data and materials

The BRFSS data used in this study are available from the Utah Department of Health and Human Services. Restrictions apply to data availability.

### Authors’ contributions

RDM helped design the study, conducted the analyses, and drafted the manuscript. BT assisted with analyses, interpretation of results, and editing of the manuscript. MM assisted with the study design including development of the study questionnaire and helped edit the manuscript. DK helped supervise analyses, interpretation of results, and editing of the manuscript. LG, MEB, and LJC, helped with study design, analyses, interpretation of results, and editing of the manuscript. JG helped design the study and edited the manuscript. PJS helped design the study and edited the manuscript. JAD helped design the study, directed and supervised its implementation, supervised analyses, and contributed to the drafting and editing of the manuscript.

### Disclaimer

Where authors are identified as personnel of the International Agency for Research on Cancer or the World Health Organization, the authors alone are responsible for the views expressed in this paper; the views do not necessarily represent the decisions, policy, or views of the International Agency for Research on Cancer or the World Health Organization.

## Acknowledgements

We wish to thank the participants in this study who dedicated their time to make this research possible. We also thank Anna Dillingham, Lynne MacLeod, MaryAnne Hunter, Lin-Marie Wright, and Shige Onda at the Utah Department of Health and Human Services whose work facilitating the BRFSS made this study possible.

## Abbreviations

BMI: body mass index
BRFSS: Behavioral Risk Factor Surveillance System
CI: confidence interval
PR: prevalence ratio
US: United States

