## Supplementary material for "Associations of tattooing with health: a population-based cross-sectional study of ∼27,000 US adults"

**Supplementary Table 1. Associations between ever receiving a tattoo and physical, oral, and mental health status among women and men: comparison of models adjusted for age only, education only, and both age and education**

|  | **Women** | | | **Men** | | |
| --- | --- | --- | --- | --- | --- | --- |
|  | **Model 1** Age only | **Model 2** Education only | **Model 3** Age + education | **Model 1** Age only | **Model 2** Education only | **Model 3** Age + education |
|  | PR (95% CI) | PR (95% CI) | PR (95% CI) | PR (95% CI) | PR (95% CI) | PR (95% CI) |
| **Physical health** | | | | | | |
| **Overall health** |  |  |  |  |  |  |
| Excellent | Ref | Ref | Ref | Ref | Ref | Ref |
| Very good | 1.16 (0.99, 1.36) | 1.01 (0.86, 1.17) | 1.12 (0.96, 1.32) | 1.04 (0.89, 1.21) | 0.98 (0.84, 1.15) | 1.03 (0.88, 1.21) |
| Good | 1.76 (1.50, 2.07) | 1.31 (1.12, 1.53) | 1.61 (1.36, 1.89) | 1.39 (1.19, 1.63) | 1.11 (0.95, 1.30) | 1.25 (1.06, 1.46) |
| Fair | 1.87 (1.51, 2.31) | 1.26 (1.02, 1.54) | 1.66 (1.34, 2.06) | 1.89 (1.51, 2.36) | 1.26 (1.01, 1.56) | 1.56 (1.24, 1.96) |
| Poor | 3.48 (2.57, 4.72) | 1.77 (1.29, 2.43) | 3.08 (2.26, 4.21) | 1.78 (1.24, 2.55) | 0.96 (0.68, 1.34) | 1.41 (0.98, 2.03) |
| **Number of days during the past 30 days when physical health was not good** | | |  |  |  |  |
| None | Ref | Ref | Ref | Ref | Ref | Ref |
| 1-7 | 0.88 (0.77, 1.00) | 1.03 (0.90, 1.17) | 0.89 (0.78, 1.02) | 0.90 (0.78, 1.04) | 0.98 (0.85, 1.13) | 0.92 (0.80, 1.07) |
| 8-14 | 1.36 (1.06, 1.75) | 1.42 (1.11, 1.81) | 1.33 (1.03, 1.72) | 1.65 (1.22, 2.22) | 1.50 (1.11, 2.01) | 1.60 (1.18, 2.16) |
| 15+ | 1.79 (1.50, 2.14) | 1.40 (1.18, 1.66) | 1.69 (1.42, 2.02) | 1.66 (1.34, 2.06) | 1.15 (0.94, 1.42) | 1.45 (1.17, 1.80) |
| **Average hours of sleep per night** | |  |  |  |  |  |
| <7 | 1.68 (1.45, 1.94) | 1.61 (1.40, 1.86) | 1.58 (1.36, 1.83) | 1.72 (1.48, 1.99) | 1.67 (1.44, 1.94) | 1.64 (1.41, 1.90) |
| 7-9 | Ref | Ref | Ref | Ref | Ref | Ref |
| >9 | 2.06 (1.44, 2.94) | 1.45 (1.02, 2.06) | 1.83 (1.26, 2.65) | 1.70 (1.07, 2.71) | 1.26 (0.81, 1.95) | 1.44 (0.91, 2.26) |
| **Body mass index (kg/m^2^)** |  |  |  |  |  |  |
| Underweight (<18.5) | 1.13 (0.74, 1.73) | 1.17 (0.76, 1.79) | 1.08 (0.70, 1.66) | 0.88 (0.55, 1.42) | 0.74 (0.46, 1.19) | 0.71 (0.44, 1.14) |
| Normal weight (18.5-24.9) | Ref | Ref^a^ | Ref | Ref | Ref^a^ | Ref |
| Overweight (25.0-29.9) | 1.21 (1.05, 1.40) | 1.00 (0.87, 1.15) | 1.17 (1.01, 1.36) | 1.08 (0.94, 1.25) | 1.00 (0.86, 1.15) | 1.12 (0.97, 1.30) |
| Obese (30+) | 1.49 (1.30, 1.71) | 1.22 (1.07, 1.40) | 1.40 (1.22, 1.61) | 1.20 (1.04, 1.39) | 1.07 (0.92, 1.24) | 1.21 (1.04, 1.40) |
| **Physical activity in past 30 days** |  |  |  |  |  |  |
| Yes | Ref | Ref^b^ | Ref | Ref | Ref^b^ | Ref |
| No | 1.33 (1.19, 1.49) | 0.97 (0.87, 1.08) | 1.21 (1.09, 1.36) | 1.78 (1.67, 1.90) | 0.98 (0.86, 1.11) | 1.09 (0.95, 1.24) |
| **Chronic pain** |  |  |  |  |  |  |
| No | Ref | Ref | Ref | Ref | Ref | Ref |
| Yes | 1.62 (1.46, 1.80) | 1.32 (1.19, 1.46) | 1.59 (1.43, 1.77) | 1.61 (1.43, 1.82) | 1.40 (1.24, 1.58) | 1.55 (1.37, 1.76) |
| **Prescription opioid use for chronic pain** | | |  |  |  |  |
| No | Ref | Ref | Ref | Ref | Ref | Ref |
| Yes | 1.69 (1.30, 2.20) | 1.20 (0.95, 1.53) | 1.62 (1.23, 2.12) | 0.99 (0.70, 1.39) | 0.67 (0.47, 0.94) | 0.91 (0.65, 1.27) |
| **Oral health** | | | | | | |
| **Visited a dentist in the past year** |  |  |  |  |  |  |
| Yes | Ref | Ref | Ref | Ref | Ref | Ref |
| No | 1.31 (1.18, 1.46) | 1.23 (1.11, 1.37) | 1.21 (1.09, 1.35) | 1.33 (1.20, 1.47) | 1.22 (1.10, 1.34) | 1.20 (1.09, 1.33) |
| **Number of permanent teeth removed due to tooth decay or gum disease** | | |  |  |  |  |
| None | Ref | Ref | Ref | Ref | Ref | Ref |
| 1-5 | 1.63 (1.38, 1.93) | 1.03 (0.88, 1.20) | 1.52 (1.29, 1.80) | 1.87 (1.57, 2.22) | 1.21 (1.02, 1.42) | 1.66 (1.40, 1.98) |
| 6 or more but not all | 2.50 (1.87, 3.34) | 0.81 (0.61, 1.08) | 2.18 (1.61, 2.96) | 3.61 (2.67, 4.89) | 1.48 (1.12, 1.96) | 2.88 (2.10, 3.95) |
| All | 3.30 (2.16, 5.04) | 0.92 (0.64, 1.31) | 2.99 (1.93, 4.61) | 3.00 (2.01, 4.48) | 0.89 (0.62, 1.27) | 2.16 (1.43, 3.26) |
| **Mental health** | | | | | | |
| **Ever had a depressive disorder** |  |  |  |  |  |  |
| No | Ref | Ref | Ref | Ref | Ref | Ref |
| Yes | 1.66 (1.55, 1.77) | 1.78 (1.67, 1.90) | 1.64 (1.53, 1.75) | 1.55 (1.39, 1.72) | 1.64 (1.47, 1.83) | 1.55 (1.39, 1.73) |
| **Number of days during the past 30 days when mental health was not good** | | |  |  |  |  |
| None | Ref | Ref | Ref | Ref | Ref | Ref |
| 1-7 | 1.04 (0.91, 1.20) | 1.46 (1.27, 1.67) | 1.08 (0.94, 1.25) | 1.03 (0.89, 1.19) | 1.27 (1.10, 1.46) | 1.09 (0.94, 1.27) |
| 8-14 | 1.52 (1.23, 1.87) | 2.15 (1.76, 2.63) | 1.51 (1.22, 1.86) | 1.25 (0.97, 1.60) | 1.43 (1.11, 1.85) | 1.23 (0.95, 1.59) |
| 15+ | 2.29 (1.97, 2.65) | 3.09 (2.68, 3.57) | 2.21 (1.91, 2.57) | 2.00 (1.69, 2.36) | 2.17 (1.83, 2.56) | 1.88 (1.59, 2.23) |
| **Within the last 30 days, how often have you felt stress?**^c^ | | |  |  |  |  |
| Never | Ref | Ref | Ref | Ref | Ref | Ref |
| Rarely | 0.90 (0.67, 1.21) | 1.05 (0.79, 1.40) | 0.91 (0.67, 1.22) | 1.01 (0.77, 1.33) | 1.15 (0.88, 1.51) | 1.06 (0.80, 1.40) |
| Sometimes | 1.06 (0.79, 1.42) | 1.35 (1.02, 1.78) | 1.03 (0.77, 1.38) | 1.62 (1.22, 2.16) | 1.91 (1.43, 2.56) | 1.72 (1.28, 2.30) |
| Usually | 1.30 (0.90, 1.87) | 2.00 (1.40, 2.86) | 1.29 (0.88, 1.87) | 1.69 (1.15, 2.47) | 1.91 (1.31, 2.81) | 1.67 (1.13, 2.47) |
| Always | 2.59 (1.65, 4.04) | 3.54 (2.29, 5.49) | 2.42 (1.54, 3.79) | 2.96 (1.90, 4.63) | 3.39 (2.14, 5.39) | 2.97 (1.87, 4.72) |

^a^BMI not included as adjustment variables in model 2

^b^Physical activity not included as an adjustment variable in model 2

^c^Stress means a situation in which a person feels tense, restless, nervous or anxious or is unable to sleep at night because their mind is troubled all the time. Within the last 30 days how often have you felt this kind of stress?

**Supplementary Table 2. Cross-sectional associations between ever receiving a tattoo and chronic health conditions among women and men**

|  | **Women** | | | | | | | **Men** | | | | | | |
| --- | --- | --- | --- | --- | --- | --- | --- | --- | --- | --- | --- | --- | --- | --- |
|  | **Never tattooed** (n=10,599) | | **Ever tattooed** (n=2,846) | | **Model 1** | **Model 2** | | **Never tattooed** (n=10,810) | | **Ever tattooed** (n=2,368) | | **Model 1** | **Model 2** | |
|  | n | (%) | n | (%) | PR (95% CI) | n | PR (95% CI) | n | (%) | n | (%) | PR (95% CI) | n | PR (95% CI) |
| **Asthma (ever)** |  |  |  |  |  |  |  |  |  |  |  |  |  |  |
| No | 8,929 | (84%) | 2,156 | (75%) | Ref |  | Ref | 9,355 | (86%) | 1,957 | (82%) | Ref |  | Ref |
| Yes | 1,616 | (16%) | 676 | (25%) | 1.53 (1.37, 1.71) | 11,672 | 1.37 (1.21, 1.55) | 1,408 | (13%) | 408 | (18%) | 1.29 (1.14, 1.47) | 12,442 | 1.26 (1.10, 1.45) |
| Unknown/missing | 54 | (0.6%) | 14 | (0.5%) |  |  |  | 47 | (0.4%) | * | (0.2%) |  |  |  |
| **Asthma (current)** |  |  |  |  |  |  |  |  |  |  |  |  |  |  |
| No | 9,291 | (88%) | 2,304 | (80%) | Ref |  | Ref | 9,855 | (91%) | 2,104 | (88%) | Ref |  | Ref |
| Yes | 1,195 | (11%) | 503 | (18%) | 1.56 (1.37, 1.78) | 11,598 | 1.38 (1.19, 1.61) | 856 | (7.7%) | 230 | (10%) | 1.35 (1.13, 1.62) | 12,363 | 1.33 (1.10, 1.62) |
| Unknown/missing | 113 | (1.1%) | 39 | (2.0%) |  |  |  | 99 | (1.0%) | 34 | (1.8%) |  |  |  |
| **COPD, emphysema, or chronic bronchitis** | |  |  |  |  |  |  |  |  |  |  |  |  |  |
| No | 9,976 | (95%) | 2,641 | (94%) | Ref |  | Ref | 10,250 | (96%) | 2,204 | (94%) | Ref |  | Ref |
| Yes | 566 | (4.4%) | 195 | (5.8%) | 1.84 (1.47, 2.31) | 11,668 | 1.05 (0.81, 1.35) | 498 | (3.7%) | 153 | (5.1%) | 1.64 (1.27, 2.11) | 12,424 | 1.18 (0.90, 1.54) |
| Unknown/missing | 57 | (0.6%) | * | (0.3%) |  |  |  | 62 | (0.5%) | 11 | (0.6%) |  |  |  |
| **Stroke** |  |  |  |  |  |  |  |  |  |  |  |  |  |  |
| No | 10,184 | (97%) | 2,762 | (98%) | Ref |  | Ref | 10,455 | (98%) | 2,292 | (97%) | Ref |  | Ref |
| Yes | 382 | (2.5%) | 81 | (2.4%) | 1.71 (1.21, 2.42) | 11,694 | 1.27 (0.89, 1.79) | 332 | (2.1%) | 72 | (2.3%) | 1.90 (1.38, 2.62) | 12,464 | 1.77 (1.26, 2.50) |
| Unknown/missing | 33 | (0.3%) | * | (<0.1%) |  |  |  | 23 | (0.2%) | * | (0.2%) |  |  |  |
| **Heart attack** |  |  |  |  |  |  |  |  |  |  |  |  |  |  |
| No | 10,244 | (97%) | 2,789 | (99%) | Ref |  | Ref | 10,115 | (95%) | 2,229 | (96%) | Ref |  | Ref |
| Yes | 310 | (2.1%) | 50 | (1.2%) | 1.25 (0.85, 1.83) | 11,684 | 0.96 (0.62, 1.49) | 640 | (4.2%) | 119 | (3.5%) | 1.39 (1.04, 1.84) | 12,426 | 1.24 (0.92, 1.66) |
| Unknown/missing | 45 | (0.4%) | * | (0.2%) |  |  |  | 55 | (0.4%) | 20 | (0.8%) |  |  |  |
| **Angina or coronary heart disease** |  |  |  |  |  |  |  |  |  |  |  |  |  |  |
| No | 10,181 | (97%) | 2,795 | (99%) | Ref |  | Ref | 10,122 | (95%) | 2,266 | (97%) | Ref |  | Ref |
| Yes | 321 | (2.2%) | 40 | (1.0%) | 0.95 (0.60, 1.49) | 11,627 | 0.81 (0.49, 1.36) | 601 | (4.0%) | 82 | (2.5%) | 1.10 (0.79, 1.54) | 12,393 | 0.96 (0.68, 1.35) |
| Unknown/missing | 97 | (0.7%) | 11 | (0.2%) |  |  |  | 87 | (0.7%) | 20 | (1.0%) |  |  |  |
| **High blood pressure (2021)** |  |  |  |  |  |  |  |  |  |  |  |  |  |  |
| No | 2,482 | (74%) | 765 | (84%) | Ref |  | Ref | 2,405 | (69%) | 556 | (65%) | Ref |  | Ref |
| Yes | 1,197 | (25%) | 211 | (16%) | 1.00 (0.97, 1.04) | 4,023 | 1.02 (0.98, 1.06) | 1,370 | (30%) | 314 | (34%) | 0.90 (0.84, 0.96) | 4,353 | 0.95 (0.89, 1.01) |
| Unknown/missing | * | (0.4%) | * | (<0.1%) |  |  |  | 19 | (0.7%) | * | (0.7%) |  |  |  |
| **High cholesterol (2021)** |  |  |  |  |  |  |  |  |  |  |  |  |  |  |
| No | 1,983 | (53%) | 551 | (57%) | Ref |  | Ref | 1,906 | (50%) | 436 | (50%) | Ref |  | Ref |
| Yes | 1,157 | (26%) | 209 | (16%) | 1.02 (0.97, 1.08) | 3,389 | 1.03 (0.97, 1.10) | 1,232 | (27%) | 219 | (22%) | 0.99 (0.93, 1.06) | 3,568 | 1.00 (0.94, 1.08) |
| Unknown/missing | 548 | (20%) | 217 | (27%) |  |  |  | 656 | (23%) | 220 | (28%) |  |  |  |
| **Kidney disease** |  |  |  |  |  |  |  |  |  |  |  |  |  |  |
| No | 10,094 | (97%) | 2,737 | (97%) | Ref |  | Ref | 10,365 | (97%) | 2,310 | (98%) | Ref |  | Ref |
| Yes | 453 | (3.1%) | 97 | (2.6%) | 1.40 (1.04, 1.88) | 11,676 | 1.44 (1.03, 2.02) | 404 | (2.7%) | 53 | (1.6%) | 1.04 (0.72, 1.49) | 12,446 | 1.05 (0.72, 1.52) |
| Unknown/missing | 52 | (0.4%) | 12 | (0.3%) |  |  |  | 41 | (0.3%) | * | (0.4%) |  |  |  |
| **Diabetes** |  |  |  |  |  |  |  |  |  |  |  |  |  |  |
| No | 9,241 | (89%) | 2,530 | (91%) | Ref |  | Ref | 9,435 | (90%) | 2,131 | (93%) | Ref |  | Ref |
| Yes, but only during pregnancy | 233 | (2.5%) | 79 | (2.9%) | 1.10 (0.78, 1.54) |  | 1.05 (0.72, 1.53) | - |  | - |  | - |  | - |
| Yes | 1,108 | (8.4%) | 233 | (6.4%) | 1.35 (1.09, 1.67) | 11,714 | 1.19 (0.93, 1.53) | 1,362 | (9.6%) | 231 | (7.0%) | 1.12 (0.94, 1.33) | 12,476 | 1.16 (0.97, 1.39) |
| Unknown/missing | 17 | (0.1%) | * | (0.2%) |  |  |  | 13 | (0.2%) | * | (0.1%) |  |  |  |
| **Arthritis, rheumatoid arthritis, gout, lupus, or fibromyalgia** | |  |  |  |  |  |  |  |  |  |  |  |  |  |
| No | 6,859 | (72%) | 2,075 | (78%) | Ref |  | Ref | 8,055 | (80%) | 1,855 | (82%) | Ref |  | Ref |
| Yes | 3,689 | (27%) | 759 | (22%) | 1.32 (1.20, 1.44) | 11,673 | 1.20 (1.09, 1.33) | 2,709 | (19%) | 496 | (17%) | 1.29 (1.15, 1.45) | 12,431 | 1.20 (1.06, 1.35) |
| Unknown/missing | 51 | (0.4%) | 12 | (0.4%) |  |  |  | 46 | (0.4%) | 17 | (0.6%) |  |  |  |
| **Non-melanoma skin cancer (2022)** |  |  |  |  |  |  |  |  |  |  |  |  |  |  |
| No | 2,733 | (92%) | 810 | (95%) | Ref |  | Ref | 2,881 | (92%) | 666 | (96%) | Ref |  | Ref |
| Yes | 353 | (7.5%) | 55 | (4.3%) | 1.28 (0.87, 1.88) | 3,454 | 1.17 (0.77, 1.77) | 347 | (7.5%) | 32 | (3.7%) | 0.94 (0.61, 1.43) | 3,687 | 0.91 (0.59, 1.40) |
| Unknown/missing | 23 | (0.6%) | * | (0.2%) |  |  |  | 29 | (0.8%) | * | (0.4%) |  |  |  |
| **Melanoma or other cancer (2022)** |  |  |  |  |  |  |  |  |  |  |  |  |  |  |
| No | 2,728 | (91%) | 777 | (92%) | Ref |  | Ref | 2,849 | (91%) | 652 | (94%) | Ref |  | Ref |
| Yes | 359 | (8.8%) | 86 | (7.8%) | 1.49 (1.07, 2.09) | 3,456 | 1.32 (0.91, 1.91) | 392 | (8.6%) | 47 | (5.4%) | 1.04 (0.70, 1.55) | 3,700 | 0.94 (0.62, 1.41) |
| Unknown/missing | 22 | (0.6%) | * | (0.4%) |  |  |  | 16 | (0.4%) | * | (0.4%) |  |  |  |

Note: n's are unweighted. Percentages are weighted and models incorporate the BRFSS weights to account for the complex survey design. For each condition, participants were asked if they had ever been diagnosed, except for asthma (current) in which participants were asked if they currently have asthma.

Model 1 is adjusted for age and education level. Model 2 is adjusted for age, education level, ever tobacco smoking, heavy drinking, BMI, and physical activity in the past 30 days.

*Censored due to cell values <11

**Supplementary Table 3. Cross-sectional associations between tattoos and cancer among women and men separately and combined**

|  | **Women** | | | | | | **Men** | | | | | | **Overall** |
| --- | --- | --- | --- | --- | --- | --- | --- | --- | --- | --- | --- | --- | --- |
|  | **Never tattooed** (n=10,599) | | **Ever tattooed** (n=2,846) | | **Model 1** | **Model 2** | **Never tattooed** (n=10,810) | | **Ever tattooed** (n=2,368) | | **Model 1** | **Model 2** | **Model 2** |
|  | n | (%) | n | (%) | PR (95% CI) | PR (95% CI) | n | (%) | n | (%) | PR (95% CI) | PR (95% CI) | PR (95% CI) |
| **Type of cancer** |  |  |  |  |  |  |  |  |  |  |  |  |  |
| None | 9,465 | (92%) | 2,575 | (93%) | Ref | Ref | 9,784 | (93%) | 2,230 | (96%) | Ref | Ref | Ref |
| Melanoma | 85 | (0.7%) | 16 | (0.5%) | 1.34 (0.65, 2.79) | 0.82 (0.37, 1.81) | 138 | (1.1%) | 15 | (0.5%) | 0.77 (0.38, 1.55) | 0.78 (0.36, 1.71) | 0.82 (0.46, 1.48) |
| Breast | 359 | (2.4%) | 67 | (1.7%) | 1.79 (1.23, 2.61) | 1.87 (1.25, 2.80) | * | (<0.1%) | - | - | - | - |  |
| Gynecologic | 145 | (1.0%) | 73 | (2.3%) | 3.19 (2.17, 4.68) | 2.28 (1.38, 3.76) | - | - | - | - | - | - | - |
| Cervical | 52 | (3.4%) | 53 | (17%) | 3.17 (2.01, 4.98) | 2.44 (1.32, 4.49) | - | - | - | - | - | - | - |
| Endometrial | 59 | (2.4%) | 12 | (2.8%) | 1.16 (0.51, 2.61) | 1.61 (0.75, 3.47) | - | - | - | - | - | - | - |
| Ovarian | 34 | (1.9%) | * | (1.9%) | 0.66 (0.27, 1.60) | 0.38 (0.15, 0.92) | - | - | - | - | - | - | - |
| Prostate | - | (0%) | - | (0%) | - | - | 316 | (1.9%) | 33 | (0.9%) | 1.18 (0.62, 2.22) | 1.15 (0.61, 2.16) | - |
| Colorectal | 51 | (0.3%) | * | (0.2%) | 1.77 (0.74, 4.22) | 1.44 (0.58, 3.56) | 46 | (0.3%) | * | (0.3%) | 2.24 (0.93, 5.42) | 1.44 (0.63, 3.29) | 1.42 (0.78, 2.61) |
| Hematologic | 58 | (0.5%) | * | (0.2%) | 0.56 (0.19, 1.63) | 0.64 (0.18, 2.21) | 71 | (0.5%) | 11 | (0.4%) | 1.06 (0.48, 2.37) | 0.67 (0.26, 1.73) | 0.66 (0.31, 1.40) |
| Thyroid | 72 | (0.6%) | 17 | (0.5%) | 0.92 (0.45, 1.89) | 1.08 (0.50, 2.37) | 21 | (0.2%) | * | (0.2%) | 0.99 (0.25, 3.97) | 1.35 (0.37, 4.93) | 1.29 (0.65, 2.54) |
| Bladder | 13 | (0.1%) | * | (<0.1%) | 1.84 (0.40, 8.55) | 1.80 (0.32, 9.95) | 35 | (0.2%) | * | (0.2%) | 2.52 (0.93, 6.81) | 2.07 (0.77, 5.56) | 2.00 (0.85, 4.68) |
| Other | 126 | (0.8%) | 33 | (0.8%) | 1.81 (1.02, 3.19) | 1.42 (0.77, 2.62) | 148 | (1.2%) | 39 | (1.1%) | 1.41 (0.88, 2.26) | 1.31 (0.78, 2.17) | 1.34 (0.91, 1.96) |
| Unknown/missing | 225 | (1.6%) | 43 | (1.2%) |  |  | 249 | (1.7%) | 20 | (0.6%) |  |  |  |

Note: n's are unweighted. Percentages are weighted and models incorporate the BRFSS weights to account for the complex survey design. Model 1 is adjusted for age and education level. Model 2 is adjusted for age, education level, ever tobacco smoking, heavy drinking, BMI, and physical activity in the past 30 days. For the overall analyses combining men and women, model 2 is additionally adjusted for sex.

**Supplementary Table 4. Cross-sectional associations between ever receiving a tattoo and chronic health conditions among women and men who never smoked**

|  | **Women** | | | | | | **Men** | | | | | |
| --- | --- | --- | --- | --- | --- | --- | --- | --- | --- | --- | --- | --- |
|  | **Never tattooed** (n=9,106) | | **Ever tattooed** (n=1,631) | | Model 1 | Model 2 | **Never tattooed** (n=8,406) | | **Ever tattooed** (n=962) | | Model 1 | Model 2 |
|  | n | (%) | n | (%) | PR (95% CI) | PR (95% CI) | n | (%) | n | (%) | PR (95% CI) | PR (95% CI) |
| **Physical health** | | | | | | | | | | | | |
| **Overall health** |  |  |  |  |  |  |  |  |  |  |  |  |
| Excellent | 1,996 | (24%) | 362 | (22%) | Ref | Ref | 2,070 | (26%) | 251 | (27%) | Ref | Ref |
| Very good | 3,498 | (38%) | 590 | (35%) | 1.07 (0.89, 1.29) | 1.04 (0.85, 1.26) | 3,167 | (38%) | 332 | (34%) | 0.95 (0.77, 1.18) | 0.94 (0.75, 1.17) |
| Good | 2,462 | (26%) | 464 | (30%) | 1.41 (1.15, 1.71) | 1.35 (1.08, 1.68) | 2,326 | (27%) | 271 | (29%) | 1.11 (0.89, 1.39) | 1.08 (0.85, 1.38) |
| Fair | 867 | (9.4%) | 163 | (11%) | 1.57 (1.19, 2.08) | 1.53 (1.12, 2.10) | 642 | (7.0%) | 88 | (8.6%) | 1.40 (1.00, 1.96) | 1.28 (0.90, 1.81) |
| Poor | 260 | (2.2%) | 48 | (2.8%) | 2.32 (1.48, 3.65) | 2.27 (1.40, 3.68) | 190 | (1.9%) | 17 | (1.2%) | 0.81 (0.42, 1.58) | 0.86 (0.43, 1.69) |
| Unknown/missing | 23 | (0.3%) | * | (0.2%) |  |  | 11 | (0.2%) | * | (0.3%) |  |  |
| **Number of days during the past 30 days when physical health was not good** |  |  |  |  |  |  |  |  |  |  |  |  |
| None | 5,441 | (58%) | 957 | (58%) | Ref | Ref | 5,743 | (68%) | 644 | (66%) | Ref | Ref |
| 1-7 | 2,153 | (26%) | 380 | (24%) | 0.79 (0.67, 0.93) | 0.77 (0.64, 0.92) | 1,766 | (22%) | 202 | (23%) | 0.96 (0.78, 1.18) | 0.96 (0.78, 1.19) |
| 8-14 | 368 | (4.4%) | 91 | (5.4%) | 1.17 (0.85, 1.60) | 1.26 (0.90, 1.76) | 242 | (2.8%) | 34 | (3.7%) | 1.42 (0.89, 2.26) | 1.28 (0.79, 2.07) |
| 15+ | 922 | (9.0%) | 177 | (11%) | 1.38 (1.08, 1.75) | 1.22 (0.93, 1.59) | 546 | (5.7%) | 69 | (6.4%) | 1.44 (1.01, 2.04) | 1.38 (0.96, 1.98) |
| Unknown/missing | 222 | (2.3%) | 26 | (1.6%) |  |  | 109 | (1.1%) | 13 | (1.1%) |  |  |
| **Average hours of sleep per night (2020 and 2022)** |  |  |  |  |  |  |  |  |  |  |  |  |
| <7 | 1,508 | (27%) | 364 | (36%) | 1.47 (1.22, 1.77) | 1.53 (1.26, 1.87) | 1,542 | (30%) | 226 | (41%) | 1.56 (1.25, 1.95) | 1.58 (1.26, 1.99) |
| 7-9 | 4,161 | (69%) | 640 | (59%) | Ref | Ref | 3,776 | (68%) | 353 | (57%) | Ref | Ref |
| >9 | 197 | (3.0%) | 38 | (3.6%) | 1.70 (1.06, 2.74) | 1.72 (1.01, 2.92) | 104 | (1.7%) | 12 | (1.5%) | 1.02 (0.49, 2.13) | 1.09 (0.51, 2.35) |
| Unknown/missing | 72 | (1.3%) | 11 | (1.0%) |  |  | 33 | (0.6%) | * | (0.4%) |  |  |
| **Body mass index (kg/m^2^)** |  |  |  |  |  |  |  |  |  |  |  |  |
| Underweight (<18.5) | 165 | (2.0%) | 28 | (2.5%) | 1.10 (0.66, 1.86) | 1.05 (0.62, 1.79) | 103 | (1.6%) | 11 | (1.1%) | 0.53 (0.25, 1.12) | 0.56 (0.27, 1.18) |
| Normal weight (18.5-24.9) | 3014 | (35%) | 567 | (35%) | Ref | Ref^a^ | 2156 | (29%) | 250 | (28%) | Ref | Ref^a^ |
| Overweight (25.0-29.9) | 2,482 | (25%) | 379 | (22%) | 1.05 (0.87, 1.26) | 1.06 (0.88, 1.28) | 3,314 | (36%) | 382 | (38%) | 1.34 (1.09, 1.64) | 1.32 (1.07, 1.63) |
| Obese (30+) | 2,333 | (25%) | 500 | (31%) | 1.39 (1.17, 1.65) | 1.41 (1.18, 1.68) | 2,550 | (29%) | 296 | (31%) | 1.31 (1.05, 1.63) | 1.29 (1.03, 1.61) |
| Unknown/missing | 1,112 | (12%) | 157 | (9.7%) |  |  | 283 | (3.9%) | 23 | (2.5%) |  |  |
| **Physical activity in past 30 days** |  |  |  |  |  |  |  |  |  |  |  |  |
| Yes | 7,462 | (83%) | 1,347 | (84%) | Ref | Ref^b^ | 7,348 | (88%) | 834 | (87%) | Ref | Ref^b^ |
| No | 1,632 | (17%) | 282 | (16%) | 1.18 (1.01, 1.37) | 1.12 (0.95, 1.32) | 1,043 | (12%) | 126 | (13%) | 1.03 (0.83, 1.29) | 1.04 (0.82, 1.31) |
| Unknown/missing | 12 | (<0.1%) | * | (<0.1%) |  |  | 15 | (0.1%) | * | (0.1%) |  |  |
| **Chronic pain (2020 and 2022)^c^** |  |  |  |  |  |  |  |  |  |  |  |  |
| No | 3,109 | (71%) | 526 | (65%) | Ref | Ref | 3,098 | (78%) | 315 | (72%) | Ref | Ref |
| Yes | 1,348 | (26%) | 261 | (30%) | 1.45 (1.26, 1.68) | 1.42 (1.21, 1.66) | 916 | (19%) | 129 | (25%) | 1.50 (1.21, 1.84) | 1.45 (1.16, 1.80) |
| Unknown/missing | 136 | (3.3%) | 30 | (4.6%) |  |  | 126 | (3.1%) | 18 | (3.5%) |  |  |
| **Prescription opioid use for chronic pain (2020 and 2022)^c^** | |  |  |  |  |  |  |  |  |  |  |  |
| No | 1,117 | (85%) | 203 | (81%) | Ref | Ref | 771 | (86%) | 119 | (95%) | Ref | Ref |
| Yes | 223 | (15%) | 55 | (18%) | 1.59 (1.10, 2.31) | 1.57 (1.07, 2.30) | 139 | (13%) | * | (4.7%) | 0.48 (0.21, 1.08) | 0.51 (0.24, 1.08) |
| Unknown/missing | * | (0.5%) | * | (0.6%) |  |  | * | (0.5%) | 0 | (0%) |  |  |
| **Oral health** | | | | | | | | | | | | |
| **Visited a dentist in the past year (2020 and 2022)** |  |  |  |  |  |  |  |  |  |  |  |  |
| Yes | 4,733 | (78%) | 786 | (74%) | Ref | Ref | 4,139 | (74%) | 407 | (69%) | Ref | Ref |
| No | 1,153 | (21%) | 256 | (25%) | 1.10 (0.94, 1.28) | 1.07 (0.91, 1.27) | 1,273 | (25%) | 184 | (30%) | 1.06 (0.90, 1.26) | 1.08 (0.91, 1.29) |
| Unknown/missing | 52 | (0.9%) | 11 | (1.0%) |  |  | 43 | (0.7%) | * | (1.0%) |  |  |
| **Number of permanent teeth removed due to tooth decay or gum disease (2020 and 2022)** |  |  |  |  |  |  |  |  |  |  |  |  |
| None | 3,725 | (69%) | 739 | (74%) | Ref | Ref | 3,619 | (73%) | 418 | (72%) | Ref | Ref |
| 1-5 | 1,604 | (23%) | 252 | (22%) | 1.28 (1.04, 1.58) | 1.28 (1.02, 1.61) | 1,426 | (22%) | 146 | (24%) | 1.55 (1.19, 2.01) | 1.47 (1.12, 1.93) |
| 6 or more but not all | 329 | (4.0%) | 36 | (2.7%) | 1.66 (1.00, 2.77) | 1.59 (0.92, 2.77) | 223 | (2.9%) | 26 | (3.8%) | 2.75 (1.55, 4.86) | 2.89 (1.61, 5.22) |
| All | 163 | (2.0%) | 16 | (0.9%) | 1.41 (0.63, 3.20) | 1.49 (0.58, 3.81) | 115 | (1.5%) | * | (0.4%) | 0.74 (0.17, 3.22) | 0.27 (0.05, 1.59) |
| Unknown/missing | 117 | (1.5%) | * | (0.8%) |  |  | 72 | (1.0%) | * | (0.5%) |  |  |
| **Mental health** | | | | | | | | | | | | |
| **Ever had a depressive disorder** |  |  |  |  |  |  |  |  |  |  |  |  |
| No | 6,863 | (73%) | 939 | (55%) | Ref | Ref | 7,233 | (84%) | 746 | (77%) | Ref | Ref |
| Yes | 2,190 | (26%) | 681 | (44%) | 1.51 (1.39, 1.65) | 1.43 (1.30, 1.56) | 1,121 | (15%) | 211 | (22%) | 1.36 (1.15, 1.61) | 1.35 (1.15, 1.60) |
| Unknown/missing | 53 | (0.8%) | 11 | (0.8%) |  |  | 52 | (0.8%) | * | (0.5%) |  |  |
| **Number of days during the past 30 days when mental health was not good** |  |  |  |  |  |  |  |  |  |  |  |  |
| None | 4,705 | (45%) | 564 | (30%) | Ref | Ref | 5,448 | (59%) | 499 | (48%) | Ref | Ref |
| 1-7 | 2,629 | (32%) | 474 | (29%) | 0.98 (0.82, 1.16) | 0.92 (0.77, 1.11) | 1,886 | (26%) | 243 | (29%) | 1.14 (0.93, 1.40) | 1.15 (0.93, 1.42) |
| 8-14 | 554 | (7.7%) | 167 | (11%) | 1.42 (1.11, 1.83) | 1.29 (0.98, 1.69) | 326 | (4.7%) | 66 | (6.4%) | 1.26 (0.88, 1.79) | 1.24 (0.86, 1.79) |
| 15+ | 1,055 | (14%) | 401 | (28%) | 2.02 (1.67, 2.44) | 1.83 (1.49, 2.24) | 646 | (9.2%) | 146 | (16%) | 1.58 (1.23, 2.05) | 1.56 (1.20, 2.03) |
| Unknown/missing | 163 | (2.1%) | 25 | (1.6%) |  |  | 100 | (1.4%) | * | (0.6%) |  |  |
| **Within the last 30 days, how often have you felt stress?^d^** **(2022)** |  |  |  |  |  |  |  |  |  |  |  |  |
| Never | 806 | (26%) | 104 | (20%) | Ref | Ref | 1,036 | (36%) | 80 | (26%) | Ref | Ref |
| Rarely | 929 | (32%) | 129 | (25%) | 0.85 (0.59, 1.23) | 0.80 (0.54, 1.18) | 820 | (33%) | 80 | (27%) | 1.05 (0.70, 1.58) | 1.02 (0.67, 1.55) |
| Sometimes | 663 | (29%) | 158 | (30%) | 0.99 (0.69, 1.41) | 0.93 (0.64, 1.36) | 475 | (20%) | 71 | (30%) | 1.88 (1.22, 2.90) | 1.80 (1.14, 2.83) |
| Usually | 212 | (9.6%) | 63 | (14%) | 1.11 (0.71, 1.74) | 0.99 (0.61, 1.58) | 150 | (7.5%) | 32 | (12%) | 1.73 (0.96, 3.10) | 1.56 (0.82, 2.98) |
| Always | 73 | (3.5%) | 44 | (11%) | 2.45 (1.41, 4.27) | 2.21 (1.17, 4.17) | 59 | (2.8%) | 16 | (5.2%) | 2.10 (0.97, 4.51) | 2.48 (1.14, 5.40) |
| Unknown/missing | * | (0.2%) | 0 | (0%) |  |  | * | (0.3%) | 0 | (0%) |  |  |

Note: n's are unweighted. Percentages are weighted and models incorporate the BRFSS weights to account for the complex survey design. Model 1 is adjusted for age and education level. Model 2 is adjusted for age, education level, heavy drinking, BMI, and physical activity in past 30 days.

*Censored due to cell values <11

^a^BMI not included as adjustment variables in model 2

^b^Physical activity not included as an adjustment variable in model 2

^c^2020: All participants (both survey legs); 2022: 50% of participants (one survey leg)

^d^Stress means a situation in which a person feels tense, restless, nervous or anxious or is unable to sleep at night because their mind is troubled all the time. Within the last 30 days how often have you felt this kind of stress?

**Supplementary Table 5. Cross-sectional associations between ever receiving a tattoo and chronic health conditions among women and men who never smoked**

|  | **Women** | | | | | | | | | | **Men** | | | | | | | |
| --- | --- | --- | --- | --- | --- | --- | --- | --- | --- | --- | --- | --- | --- | --- | --- | --- | --- | --- |
|  | **Never tattooed** | | | **Ever tattooed** | | | Model 1 | | Model 2 | | **Never tattooed** | | **Ever tattooed** | | Model 1 | | Model 2 | |
|  | (n=9,106) | | | (n=1,631) | | |  |  |  |  | (n=8,406) | | (n=962) | |  |  |  |  |
|  | n | (%) | n | | (%) | PR (95% CI) | | PR (95% CI) | | n | | (%) | n | (%) | | PR (95% CI) | | PR (95% CI) |
| **Asthma (ever)** |  |  |  | |  |  | |  | |  | |  |  |  | |  | |  |
| No | 7,747 | (85%) | 1,257 | | (76%) | Ref | | Ref | | 7,269 | | (86%) | 802 | (82%) | | Ref | | Ref |
| Yes | 1,313 | (15%) | 369 | | (24%) | 1.55 (1.35, 1.78) | | 1.45 (1.25, 1.68) | | 1,101 | | (14%) | 160 | (18%) | | 1.25 (1.04, 1.50) | | 1.26 (1.04, 1.53) |
| Unknown/missing | 46 | (0.6%) | * | | (0.2%) |  | |  | | 36 | | (0.5%) | 0 | (0%) | |  | |  |
| **Asthma (current)** |  |  |  | |  |  | |  | |  | |  |  |  | |  | |  |
| No | 8,035 | (88%) | 1,337 | | (81%) | Ref | | Ref | | 7,661 | | (91%) | 866 | (89%) | | Ref | | Ref |
| Yes | 973 | (11%) | 276 | | (18%) | 1.59 (1.35, 1.88) | | 1.46 (1.23, 1.75) | | 667 | | (7.7%) | 86 | (9.7%) | | 1.28 (0.99, 1.662) | | 1.31 (1.00, 1.71) |
| Unknown/missing | 98 | (1.1%) | 18 | | (1.5%) |  | |  | | 78 | | (1.0%) | * | (1.1%) | |  | |  |
| **COPD, emphysema, or chronic bronchitis** | |  |  | |  |  | |  | |  | |  |  |  | |  | |  |
| No | 8,743 | (97%) | 1,573 | | (97%) | Ref | | Ref | | 8,137 | | (97%) | 932 | (97%) | | Ref | | Ref |
| Yes | 316 | (2.9%) | 52 | | (3.0%) | 1.39 (0.97, 2.00) | | 1.23 (0.82, 1.83) | | 224 | | (2.2%) | 25 | (2.3%) | | 1.18 (0.70, 1.97) | | 1.27 (0.75, 2.15) |
| Unknown/missing | 47 | (0.5%) | * | | (0.3%) |  | |  | | 45 | | (0.5%) | * | (0.9%) | |  | |  |
| **Stroke** |  |  |  | |  |  | |  | |  | |  |  |  | |  | |  |
| No | 8,814 | (98%) | 1,598 | | (99%) | Ref | | Ref | | 8,182 | | (98%) | 942 | (98%) | | Ref | | Ref |
| Yes | 269 | (2.0%) | 33 | | (1.3%) | 1.30 (0.80, 2.13) | | 1.15 (0.69, 1.92) | | 208 | | (1.7%) | 18 | (1.2%) | | 1.40 (0.73, 2.70) | | 1.58 (0.81, 3.09) |
| Unknown/missing | 23 | (0.3%) | 0 | | (0%) |  | |  | | 16 | | (0.2%) | * | (0.5%) | |  | |  |
| **Heart attack** |  |  |  | |  |  | |  | |  | |  |  |  | |  | |  |
| No | 8,845 | (98%) | 1,613 | | (99%) | Ref | | Ref | | 7,952 | | (96%) | 923 | (97%) | | Ref | | Ref |
| Yes | 225 | (1.7%) | 18 | | (0.8%) | 1.14 (0.63, 2.06) | | 0.87 (0.45, 1.67) | | 417 | | (3.3%) | 34 | (2.2%) | | 1.35 (0.86, 2.11) | | 1.47 (0.93, 2.32) |
| Unknown/missing | 36 | (0.4%) | 0 | | (0%) |  | |  | | 37 | | (0.4%) | * | (0.5%) | |  | |  |
| **Angina or coronary heart disease** |  |  |  | |  |  | |  | |  | |  |  |  | |  | |  |
| No | 8,795 | (98%) | 1,609 | | (99%) | Ref | | Ref | | 7,968 | | (96%) | 933 | (98%) | | Ref | | Ref |
| Yes | 237 | (1.9%) | 18 | | (0.9%) | 1.05 (0.58, 1.92) | | 1.06 (0.56, 1.98) | | 383 | | (3.2%) | 23 | (1.4%) | | 0.86 (0.49, 1.49) | | 0.90 (0.52, 1.58) |
| Unknown/missing | 74 | (0.6%) | * | | (0.3%) |  | |  | | 55 | | (0.5%) | * | (0.7%) | |  | |  |
| **High blood pressure (2021)** |  |  |  | |  |  | |  | |  | |  |  |  | |  | |  |
| No | 2,201 | (76%) | 483 | | (87%) | Ref | | Ref | | 1,972 | | (72%) | 257 | (73%) | | Ref | | Ref |
| Yes | 961 | (23%) | 95 | | (13%) | 1.01 (0.97, 1.05) | | 1.01 (0.97, 1.05) | | 964 | | (27%) | 106 | (26%) | | 0.95 (0.89, 1.03) | | 0.98 (0.91, 1.05) |
| Unknown/missing | * | (0.3%) | 0 | | (0%) |  | |  | | 15 | | (0.7%) | * | (0.6%) | |  | |  |
| **High cholesterol (2021)** |  |  |  | |  |  | |  | |  | |  |  |  | |  | |  |
| No | 1,735 | (54%) | 342 | | (58%) | Ref | | Ref | | 1,500 | | (50%) | 189 | (54%) | | Ref | | Ref |
| Yes | 952 | (25%) | 101 | | (13%) | 1.03 (0.97, 1.09) | | 1.03 (0.97, 1.10) | | 924 | | (26%) | 82 | (19%) | | 1.02 (0.94, 1.12) | | 1.03 (0.94, 1.12) |
| Unknown/missing | 481 | (21%) | 135 | | (29%) |  | |  | | 527 | | (24%) | 94 | (27%) | |  | |  |
| **Kidney disease** |  |  |  | |  |  | |  | |  | |  |  |  | |  | |  |
| No | 8,696 | (97%) | 1,572 | | (97%) | Ref | | Ref | | 8,062 | | (97%) | 941 | (98%) | | Ref | | Ref |
| Yes | 368 | (2.9%) | 54 | | (2.7%) | 1.77 (1.24, 2.53) | | 1.74 (1.20, 2.52) | | 313 | | (2.6%) | 19 | (1.1%) | | 0.79 (0.44, 1.42) | | 0.88 (0.49, 1.57) |
| Unknown/missing | 42 | (0.4%) | * | | (0.2%) |  | |  | | 31 | | (0.4%) | * | (0.6%) | |  | |  |
| **Diabetes** |  |  |  | |  |  | |  | |  | |  |  |  | |  | |  |
| No | 7,988 | (90%) | 1,477 | | (92%) | Ref | | Ref | | 7,410 | | (91%) | 892 | (95%) | | Ref | | Ref |
| Yes, but only during pregnancy | 199 | (2.5%) | 44 | | (2.8%) | 1.08 (0.72, 1.64) | | 1.09 (0.69, 1.73) | | - | | - | - | - | | - | | - |
| Yes | 905 | (7.6%) | 108 | | (5.1%) | 1.32 (1.00, 1.73) | | 1.32 (0.98, 1.79) | | 987 | | (8.8%) | 68 | (5.1%) | | 0.99 (0.73, 1.33) | | 1.05 (0.78, 1.41) |
| Unknown/missing | 14 | (0.1%) | * | | (0.1%) |  | |  | | * | | (0.2%) | * | (0.2%) | |  | |  |
| **Arthritis, rheumatoid arthritis, gout, lupus, or fibromyalgia** | | | | | |  | |  | |  | |  |  |  | |  | |  |
| No | 6,098 | (75%) | 1,285 | | (84%) | Ref | | Ref | | 6,487 | | (83%) | 807 | (85%) | | Ref | | Ref |
| Yes | 2,965 | (25%) | 341 | | (16%) | 1.14 (1.00, 1.29) | | 1.18 (1.04, 1.35) | | 1,882 | | (17%) | 147 | (14%) | | 1.30 (1.07, 1.59) | | 1.32 (1.08, 1.60) |
| Unknown/missing | 43 | (0.4%) | * | | (0.4%) |  | |  | | 37 | | (0.4%) | * | (0.8%) | |  | |  |
| **Non-melanoma skin cancer (2022)** |  |  |  | |  |  | |  | |  | |  |  |  | |  | |  |
| No | 2,367 | (92%) | 468 | | (96%) | Ref | | Ref | | 2,277 | | (93%) | 266 | (97%) | | Ref | | Ref |
| Yes | 307 | (7.6%) | 29 | | (4.1%) | 1.17 (0.71, 1.92) | | 0.97 (0.60, 1.59) | | 249 | | (6.7%) | 11 | (2.6%) | | 0.87 (0.44, 1.73) | | 0.92 (0.46, 1.82) |
| Unknown/missing | 17 | (0.6%) | * | | (0.2%) |  | |  | | 22 | | (0.8%) | * | (0.5%) | |  | |  |
| **Melanoma or other cancer (2022)** |  |  |  | |  |  | |  | |  | |  |  |  | |  | |  |
| No | 2,378 | (91%) | 452 | | (92%) | Ref | | Ref | | 2,254 | | (92%) | 264 | (97%) | | Ref | | Ref |
| Yes | 295 | (8.1%) | 44 | | (7.2%) | 1.56 (1.00, 2.43) | | 1.48 (0.95, 2.31) | | 283 | | (7.9%) | 15 | (3.0%) | | 0.83 (0.40, 1.70) | | 0.75 (0.34, 1.65) |
| Unknown/missing | 18 | (0.7%) | * | | (0.5%) |  | |  | | 11 | | (0.5%) | 0 | (0%) | |  | |  |

Note: n's are unweighted. Percentages are weighted and models incorporate the BRFSS weights to account for the complex survey design. For each condition, participants were asked if they had ever been diagnosed, except for asthma (now) in which participants were asked if they currently have asthma.

Model 1 is adjusted for age and education level. Model 2 is adjusted for age, education level, heavy drinking, BMI, and physical activity in the past 30 days.

*Censored due to cell values <11

**Supplementary Table 6. Cross sectional associations between tattoos and cancer among women and men who never smoked separately and combined**

|  | **Women** | | | | | | **Men** | | | | | | **Overall** |
| --- | --- | --- | --- | --- | --- | --- | --- | --- | --- | --- | --- | --- | --- |
|  | **Never tattooed** | | **Ever tattooed** | | Model 1 | Model 2 | **Never tattooed** | | **Ever tattooed** | | Model 1 | Model 2 | Model 2 |
|  | (n=9,106) | | (n=1,631) | |  |  | (n=8,406) | | (n=962) | |  |  |  |
|  | n | (%) | n | (%) | PR (95% CI) | PR (95% CI) | n | (%) | n | (%) | PR (95% CI) | PR (95% CI) | PR (95% CI) |
| **Type of cancer** |  |  |  |  |  |  |  |  |  |  |  |  |  |
| None | 8,195 | (94%) | 1,501 | (95%) | Ref | Ref | 7,683 | (95%) | 914 | (97%) | Ref | Ref | Ref |
| Melanoma | 69 | (0.6%) | * | (0.5%) | 1.26 (0.45, 3.58) | 0.96 (0.43, 2.16) | 104 | (1.0%) | * | (0.5%) | 1.02 (0.33, 3.13) | 1.15 (0.37, 3.58) | 1.18 (0.59, 2.35) |
| Breast | 295 | (2.4%) | 44 | (1.7%) | 1.93 (1.23, 3.02) | 1.84 (1.14, 2.96) | * | (<0.1%) | 0 | (0%) | - | - | - |
| Gynecologic | 106 | (0.8%) | 24 | (1.3%) | 2.81 (1.46, 5.39) | 3.57 (1.82, 7.00) | - | | - | | - | - | - |
| Cervical | 33 | (2.2%) | 18 | (13%) | 4.19 (2.03, 8.65) | 4.45 (2.10, 9.44) | - | | - | | - | - | - |
| Endometrial | 50 | (2.7%) | * | (1.7%) | 0.66 (0.18, 2.49) | 0.95 (0.26, 3.41) | - | | - | | - | - | - |
| Ovarian | 26 | (1.4%) | * | (0.4%) | 0.23 (0.05, 1.19) | 0.42 (0.08, 2.20) | - | | - | | - | - | - |
| Prostate | - | | - | | - | - | 220 | (1.7%) | 16 | (0.7%) | 1.22 (0.57, 2.59) | 1.27 (0.60, 2.71) | - |
| Colorectal | 38 | (0.3%) | * | (<0.1%) | 0.16 (0.02, 1.23) | 0.21 (0.03, 1.61) | 34 | 0.3%) | * | (<0.1%) | 0.56 (0.07, 4.29) | 0.58 (0.08, 4.46) | 0.36 (0.18, 7.06) |
| Hematologic | 51 | (0.5%) | * | (0.3%) | 0.72 (0.21, 2.53) | 0.78 (0.19, 3.24) | 48 | (0.5%) | * | (0.4%) | 1.21 (0.37, 4.01) | 0.80 (0.21, 2.98) | 0.83 (0.32, 2.17) |
| Thyroid | 63 | (0.6%) | 11 | (0.5%) | 0.92 (0.38, 2.26) | 1.15 (0.46, 2.87) | 16 | (0.2%) | * | (0.2%) | 0.92 (0.15, 5.69) | 0.86 (0.13, 5.54) | 1.08 (0.47, 2.51) |
| Bladder | * | (<0.1%) | * | (<0.1%) | 2.42 (0.43, 13.52) | 2.56 (0.47, 13.85) | 19 | (0.1%) | 0 | (0%) | - | - | 1.41 (0.31, 6.49) |
| Other | 97 | (0.7%) | 15 | (0.6%) | 1.59 (0.76, 3.32) | 1.53 (0.69, 3.38) | 102 | (1.0%) | 13 | (0.8%) | 1.15 (0.51, 2.61) | 1.24 (0.55, 2.80) | 1.35 (0.08, 2.38) |
| Unknown/missing | 182 | (1.6%) |  |  |  |  | 178 | (1.5%) | * | (0.4%) |  |  |  |

Note: n's are unweighted. Percentages are and models incorporate the BRFSS weights to account for the complex survey design. For each condition, participants were asked if they had ever been diagnosed, except for asthma (now) in which participants were asked if they currently have asthma.

Model 1 is adjusted for age and education level. Model 2 is adjusted for age, education level, heavy drinking, BMI, physical activity in past 30 days. For the overall analyses combining men and women, model 2 is additionally adjusted for sex.

*Censored due to cell values <11
